# Host-pathogen dynamics in longitudinal clinical specimens from patients with COVID-19

**DOI:** 10.1101/2021.04.27.21256149

**Authors:** Michelle J. Lin, Victoria M. Rachleff, Hong Xie, Lasata Shrestha, Nicole A.P. Lieberman, Vikas Peddu, Amin Addetia, Amanda M. Casto, Nathan Breit, Patrick C. Mathias, Meei-Li Huang, Keith R. Jerome, Alexander L. Greninger, Pavitra Roychoudhury

## Abstract

**Background:** Rapid dissemination of SARS-CoV-2 sequencing data to public repositories has enabled widespread study of viral genomes, but studies of longitudinal specimens from infected persons are relatively limited. Analysis of longitudinal specimens enables understanding of how host immune pressures drive viral evolution *in vivo*.

**Methods and findings:** Here we performed sequencing of 49 longitudinal SARS-CoV-2-positive samples from 20 patients in Washington State collected between March and September of 2020. Viral loads declined over time with an average increase in RT-PCR cycle threshold (Ct) of 0.87 per day. We found that there was negligible change in SARS-CoV-2 consensus sequences over time, but identified a number of nonsynonymous variants at low frequencies across the genome. We observed enrichment for a relatively small number of these variants, all of which are now seen in consensus genomes across the globe at low prevalence. In one patient, we saw rapid emergence of various low-level deletion variants at the N-terminal domain of the spike glycoprotein, some of which have previously been shown to be associated with reduced neutralization potency from sera. In a subset of samples that were sequenced using metagenomic methods, differential gene expression analysis showed a downregulation of cytoskeletal genes that was consistent with a loss of ciliated epithelium during infection and recovery. We also identified co-occurrence of bacterial species in samples from multiple hospitalized individuals.

**Conclusions:** These results demonstrate that the intrahost genetic composition of SARS-CoV-2 is dynamic during the course of COVID-19, and highlight the need for continued surveillance and deep sequencing of minor variants.

## Introduction

SARS-CoV-2 is the cause of coronavirus disease 2019 (COVID-19). There have been over 138 million COVID-19 cases and over 2.9 million total deaths due to COVID-19 worldwide. Genomic analyses of longitudinal specimens within infected persons are critical to understanding the evolutionary trajectory of SARS-CoV-2. Sequencing of longitudinal samples from infected individuals allows examination of viral genetic diversity, host immune response, and dynamics of co-infecting pathogens over the course of infection and recovery. Within-host variants arise during viral replication and a number of processes shape their frequencies over time. These include selective pressures at different scales (molecular, immunological, epidemiological), host heterogeneity, spatial structure, population bottlenecks, and other stochastic processes [1]. Within-host variants may impact the success of vaccines and therapeutics, and a fraction of variants that arise will be transmitted between hosts and can eventually reach fixation in the population. Recent studies of within-host diversity of SARS-CoV-2 have demonstrated the presence of low levels of minor variants and infrequent emergence of escape mutations [2–6]. Of particular note, deletions in the N-terminal domain of the spike glycoprotein have been observed in chronically infected immunocompromised patients that are associated with SARS-CoV-2 escape from sera [7–10], and are present in current circulating lineages of concern.

Here we examined longitudinal clinical specimens collected from 20 COVID-19-positive patients in Washington State. With metagenomic sequencing we identified changes in host gene expression and bacterial co-occurrences, which may be associated with recovery. We found negligible change in viral consensus sequences over time, but detectable changes in variant allele frequencies that are only weakly predictive of future consensus changes across the globe. We further observed rapid emergence of deletion variants in the N-terminus domain of the spike glycoprotein in one patient, potentially suggesting within-host SARS-CoV-2 evasion of NTD-directed antibodies. Taken together our results support the limited emergence and fixation of escape variants during a typical infection, and also highlight the need to monitor minor variants due to their potential impact on vaccine and therapeutic efficacy.

## Methods

### Sample collection and clinical testing for SARS-CoV-2

Specimens were obtained as part of clinical testing for SARS-CoV-2 ordered by local healthcare providers or collected at drive-through testing sites. RNA was extracted and the presence of SARS-CoV-2 was detected by RT-PCR as previously described using either the emergency use-authorized UW CDC-based laboratory-developed test, Hologic Panther Fusion or Roche cobas SARS-CoV-2 tests [11,12].

### Chart review

We reviewed clinical records of patients who received care within the UW network under University of Washington IRB: STUDY00000408. Information obtained from medical records included sex, age, comorbidities, medication, hospital or critical care admission, and discharge status.

### Sequencing and bioinformatic analysis

Sequencing was attempted on all samples with a positive RT-PCR assay result that had a Ct ≤36 using either a metagenomic approach described previously [13] via IDT probe-capture [14], or using Swift Biosciences’ Normalase Amplicon Panel library preparation [15]. Libraries were sequenced on Illumina MiSeq, NextSeq, or NovaSeq instruments using 300, 150, 100, or 75 bp reads. Consensus sequences were assembled using TAYLOR [15], a custom bioinformatics pipeline (https://github.com/greninger-lab/covid_swift_pipeline) with or without an additional primer clipping step depending on library preparation method. Consensus sequences were deposited to Genbank and GISAID [16], and raw reads to SRA under Bioproject PRJNA610428. Accessions are available in Supplementary Table 1.

Consensus sequences from each individual were aligned with the reference sequence NC_045512 using MAFFT v7 [17]. Clade assignments were generated using Pangolin (http://github.com/cov-lineages/pangolin) and Nextstrain [18] in December 2020. Consensus sequences with <5% Ns across the length of the genome were considered for further analysis.

Variants were also called with TAYLOR from aligned reads. Variants leading to coding changes with a sequencing depth of >100 and an allele frequency >0.01 were subjected to further analysis. We excluded mutations in the first 100 and last 50 bases, as well as variants determined to be due to sequencing error. Most samples were re-prepped and sequenced multiple times to ensure accuracy of variant calls. Variants at positions 6700, 11081-83, 19989, and 29056 were observed in a large number of samples but were determined to be the result of homopolymer sequencing error and were excluded.

### RNAseq analysis

Reads were adapter and quality trimmed with Trimmomatic v0.39 [19] using the call “leading 3 trailing 3 slidingwindow:4:15 minlen 20”, then pseudoaligned to the hg38-derived human transcriptome using Kallisto v0.46 [20]. Only samples with more than 900,000 reads pseudo-aligned to the human genome were used for analysis. Differential expression analysis using the Wald test was performed using DEseq2 [21] and deemed significant at a Benjamini-Hochberg adjusted p value < 0.1. Statistical enrichment of Gene Ontology Biological Processes was performed on all significant genes using the R package clusterProfiler [22]. Raw counts have been submitted to the Gene Expression Omnibus, accession GSE173310.

### Metagenomic analysis

Raw FASTQ files were analyzed using CLOMP v0.1.4 (https://github.com/FredHutch/CLOMP) as previously described [23]. Samples with more than 10 million reads were randomly down-sampled to 10 million reads before analysis using the “sample” command in seqtk (https://github.com/lh3/seqtk). The pipeline output was visualized using the Pavian metagenomic explorer [24], and reads per million (RPM) calculations were done using a custom R script. Results were filtered to highlight RPM counts for a shortlist of clinically relevant taxa (S4 Table). Samples were determined to be positive if the species level RPM was at least 30 for viruses, and 100 for bacteria.

## Results

### Sequencing longitudinal samples from SARS-CoV2 infected individuals

Residual clinical specimens were obtained from the University of Washington (UW) Virology Lab after testing for SARS-CoV-2 [25]. By reviewing our laboratory information system, we identified 20 individuals who had two or more positive or inconclusive samples collected between March and September 2020 (Table 1 and S1 Table). Inconclusive samples had at least one PCR target detected. A majority of samples came from inpatients who received care within the UW Medicine system, which includes UW Medical Center, Harborview Medical Center, and Northwest Hospital. Samples came from individuals with a mean age of 70 (range 42 – 99), many with severe disease given the availability of multiple samples from these patients. Consistent with other reports [26], we observed that viral load declined over time in most patients with two or more positive or inconclusive samples with an average increase in RT-PCR cycle threshold (Ct) of 0.87 per day (Fig 1 and S1 Fig).

**Table 1.** Demographics and clinical characteristics of patients included in study.

| Characteristics | (N = 20) |
| --- | --- |
| **Mean age, y (SD)** | 70 (18) |
| **Male, n (%)** | 13 (65) |
| **Race, n (%)** |  |
| White | 12 (60) |
| Asian | 4 (15) |
| Black or African American | 2 (10) |
| American Indian or Alaska Native | 1 (5) |
| Unknown or Unavailable | 1 (5) |
| **Comorbidities, n (%)** |  |
| Hypertension | 10 (50) |
| Diabetes | 7 (35) |
| Obesity | 2 (10) |
| Asthma | 1 (5) |
| **Treatment, n (%)** |  |
| Convalescent Plasma | 2 (10) |
| Hydroxychloroquine | 2 (10) |
| Azithromycin | 5 (25) |
| Tocilizumab | 1 (5) |
| ACTT-1 Trial | 2 (10) |
| No Treatment | 8 (40) |
| Unknown | 4 (20) |
| **Hospital outcomes, n (%)** |  |
| Hospital admission | 14 (70) |
| ICU admission for COVID-19 | 4 (20) |
| Survival to discharge | 18 (90) |

Different categories (in bold) and their subcategories are shown in the first column, with their respective number of patients in the second column. In parentheses, standard deviation is indicated in the first row, and percentages for all other rows.

**Fig 1.**
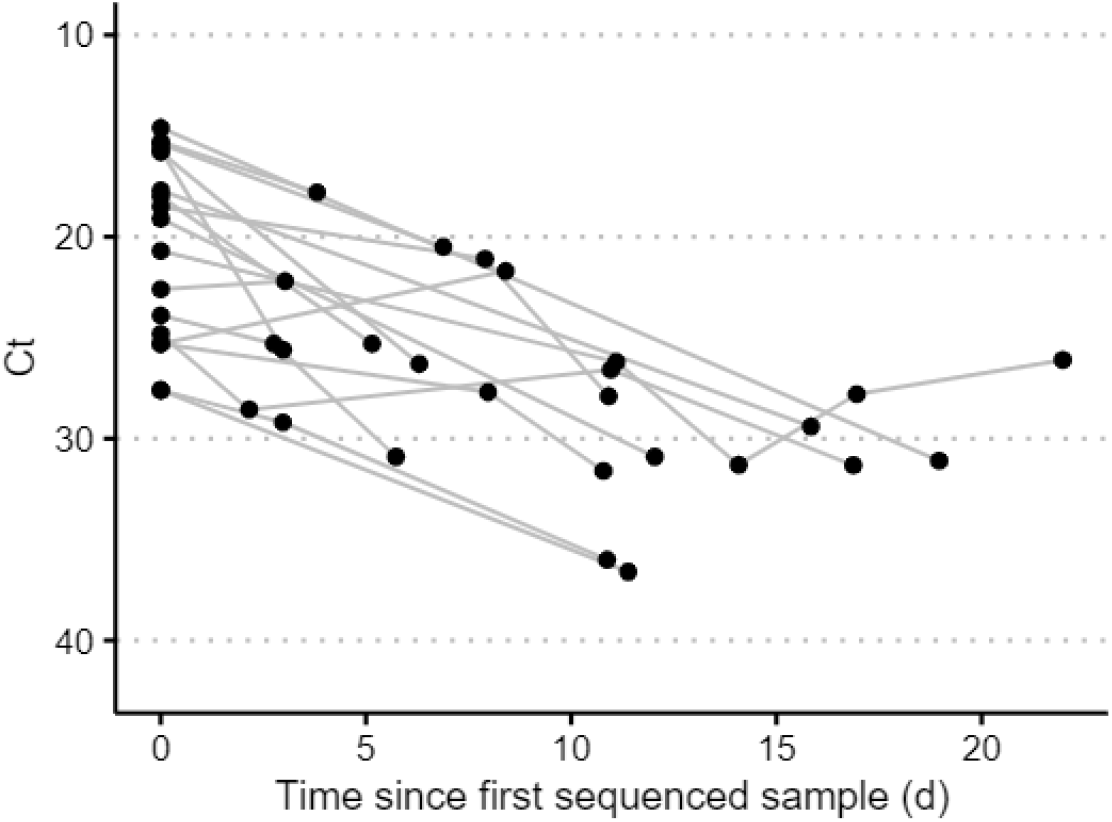
Viral load dynamics in sequenced samples.

Dots represent a unique sequenced sample. Lines connect samples from a single patient. Same day samples are not shown (see S1 Fig).

A total of 49 samples (47 nasopharyngeal and 2 oropharyngeal swabs) were sequenced with sufficient reads to be included in this study (S2 Fig). Ct values for these samples ranged between 14.6 and 36.6. The length of time between collection dates for sequenced samples from the same individual ranged from 0 to 22 days. Samples collected on the same date were sequenced for three individuals. We sequenced nasopharyngeal and oropharyngeal samples from P004 collected at the time of autopsy, two samples from P006 collected at the same time during a hospital admission, and samples collected 9 hours apart from P012 during an emergency room visit.

### Negligible change in consensus sequences over time

We obtained full-length viral genome sequences with less than 2% unknown bases (Ns) for two or more time points in 14 out of 20 individuals, plus an additional two individuals with paired nasopharyngeal and oropharyngeal swabs collected at the same timepoint (Table 2, n = 38 sequences). After masking ambiguous sites and regions with sequencing or assembly errors, we found no differences between the first and subsequent consensus sequences in 15 out of 16 patients. In one patient (P001), two samples collected 3 days apart had 4 differences between their consensus sequences at reference positions 15418 (G/T), 26262 (G/T), 27899 (T/A), and 27944 (T/C). Two of these differences lead to coding changes (A660S in nsp12 and Q2K in ORF8). Variant alleles were observed for all four positions at low frequencies in the earlier sample.

**Table 2:** Consensus sequence analysis of SARS-CoV-2 in longitudinal specimens.

| Patient | Sample # | Days since symptom onset | Ct Value | %Ns | Clade (Nextclade/Pangolin) | Number of nt differences relative to first sample |
| --- | --- | --- | --- | --- | --- | --- |
| P001 | 1 | Asymptomatic | 22.6 | 0.0% | 19B/A.1 | - |
|  | 2 | 3** | 22.2 | 0.0% |  | 4 |
| P003 | 1 | Unknown | 18.0 | 0.0% | 19B/A.1 | - |
|  | 2 | 5** | 25.3 | 0.0% |  | 0 |
| P005 | 1 | 0* | 19.1 | 0.3% | 19B/A.1 | - |
|  | 2 | 12 | 30.9 | 0.0% |  | 0 |
| P006 | 1 | Asymptomatic | 25.6 | 0.0% | 19B/A.1 | - |
|  | 2 | 0** | 29.7 | 0.0% |  | 0 |
| P007 | 1 | 0* | 15.8 | 0.0% | 19B/A.1 | - |
|  | 2 | 6 | 26.3 | 0.0% |  | 0 |
| P008 | 1 | 0* | 17.7 | 0.0% | 19B/A.1 | - |
|  | 2 | 16 | 29.4 | 0.0% |  | 0 |
| P009 | 1 | 0* | 25.3 | 0.0% | 20C/B.1.21 | - |
|  | 2 | 9 | 21.7 | 0.0% |  | 0 |
| P010 | 1 | -7 | 20.7 | 0.0% | 19B/A.1 | - |
|  | 2 | 4 | 26.2 | 0.0% |  | 0 |
|  | 3 | 7 | 31.3 | 0.0% |  | 0 |
|  | 4 | 15 | 26.1 | 0.0% |  | 0 |
| P011 | 1 | 0 | 25.3 | 0.5% | 20C/B.1.21 | - |
|  | 2 | 8 | 27.7 | 0.0% | 20C/B.1.21 | 0 |
|  | 3 | 11 | 31.6 | 0.1% |  | 0 |
| P012 | 1 | 5 | 21.6 | 0.7% | 20C/B.1.21 | - |
|  | 2 | 5 | 19.8 | 0.0% |  | 0 |
| P014 | 1 | Asymptomatic | 27.6 | 0.0% | 19B/A.1 | - |
|  | 2 | 3** | 29.2 | 1.6% |  | 0 |
| P015 | 1 | 3 | 18.5 | 0.0% | 19B/A.1 | - |
|  | 2 | 11 | 21.1 | 0.0% |  | 0 |
|  | 3 | 14 | 27.9 | 0.0% |  | 0 |
| P016 | 1 | 16 | 23.9 | 0.0% | 20C/B.1 | - |
|  | 2 | 19 | 25.3 | 0.0% |  | 0 |
|  | 3 | 22 | 30.9 | 0.0% |  | 0 |
| P017 | 1 | 10 | 24.8 | 0.0% | 19B/A.1 | - |
|  | 2 | 13 | 28.6 | 0.0% |  | 0 |
|  | 3 | 21 | 26.6 | 0.0% |  | 0 |
| P018 | 1 | Unknown | 15.3 | 0.0% | 20B/B.1.1.77 | - |
|  | 2 | 3** | 17.8 | 0.0% |  | 0 |
| P019 | 1 | Unknown | 14.6 | 0.0% | 20A/B.1 | - |
|  | 2 | 19** | 31.1 | 0.3% |  | 0 |

All patients with less than 2% unknown bases (Ns) are included. The last column indicates nucleotide differences compared to the first sample collected for each respective patient. One asterisk (\*) indicates symptoms were present at first time point but exact date of symptom onset is unknown. Two asterisks (\*\*) indicate days since first sample.

### Low frequency variants detected across the genome

We analyzed intrahost viral genetic variation by examining all sites with >100x locus depth, masking known problematic sites (see Methods). We examined sites in 47 samples from 20 different patients and found a total of 1267 unique non-synonymous variants relative to the Wuhan-Hu-1 (NC_045512.2) reference genome present at frequencies between 5-95% (Fig 2). nsp3 had the highest number of variant sites (286, 22.57%), followed by the spike glycoprotein (186, 14.68%) and nsp12 (133, 10.50%). When adjusted for gene length, mutations are most prevalent in nsp2 (103, 8.13%), at 0.053 variant sites per nucleotide. Variant frequencies were reproducible across replicates (S2 Fig).

**Fig 2.**
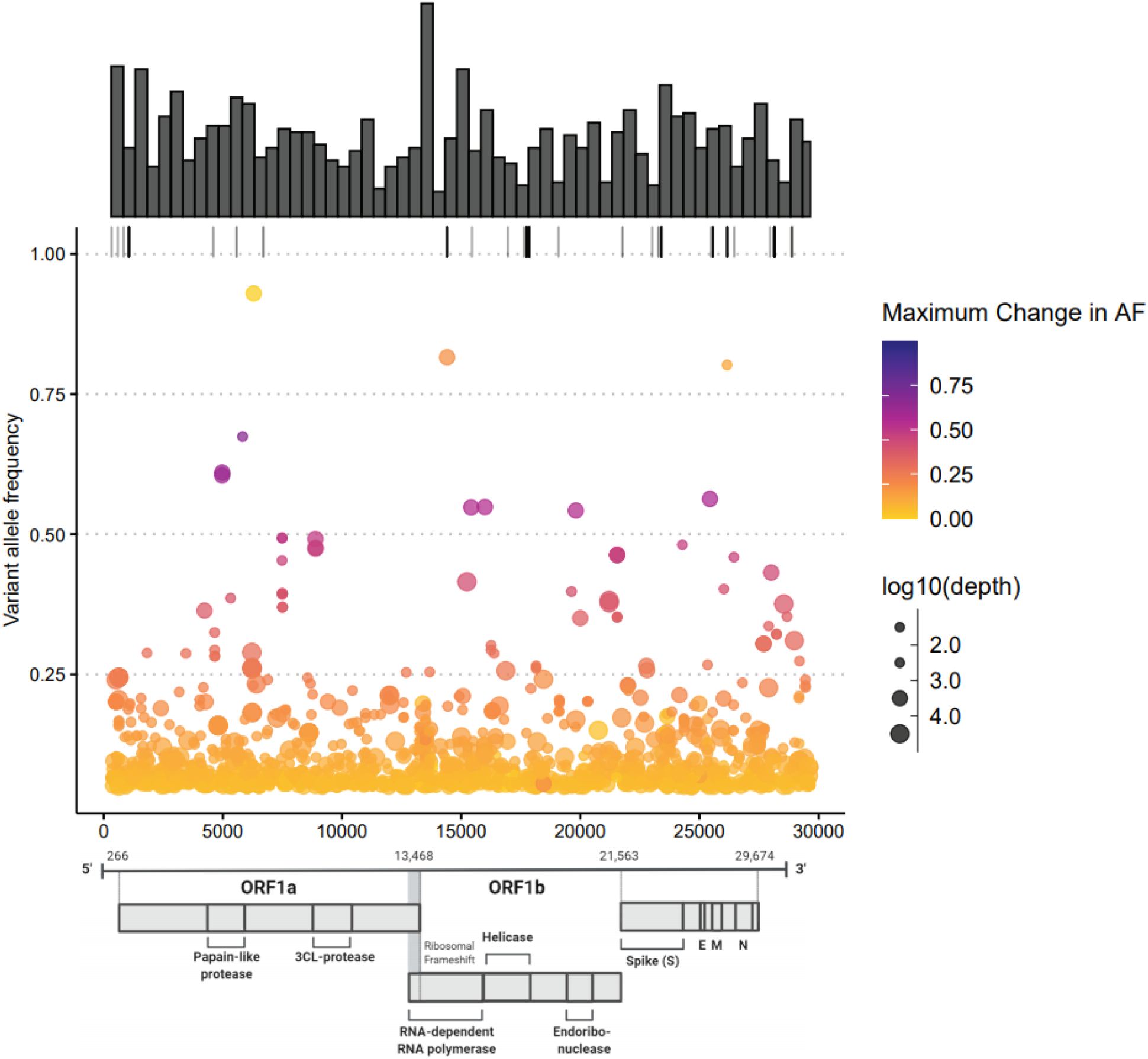
Low frequency variation is abundant but only a small number of variants exhibit a significant change in allele frequency over the course of infection.

Each dot represents a coding change in a single sample relative to the Wuhan-Hu-1 (NC_045512.2) reference genome with variant allele frequency between 5-95% and at least 100x coverage at the site. Color scale represents the change in allele frequency across time points in the same patient with darker colors representing variants that had greater changes in frequency across samples. Small dark grey marks along the top margin shows positions with variant frequencies >95% (fixed mutations relative to the reference). Size of circles indicates sequencing depth at the site. Marginal histogram shows distribution of variants using bin width of 500 nt.

Of the seven most commonly observed variants in our dataset (Table 3), the three most frequent define the Washington state outbreak clade [28,29] and the rest of the variants are clade-defining mutations in Nextstrain clades 20A and 20C. Nine out of 20 patients had the spike protein mutation D614G (A23403G), which has been associated with increased transmissibility and higher viral loads [30,31]. While this variant was rare at the beginning of the pandemic, it reached near fixation in the global SARS-CoV-2 population by June 2020 [32]. This rapid rise in prevalence is reflected in our data, as this D614G mutation is present in all three patients with samples collected during or after June 2020. In 10 out of the 11 patients with the 614D variant, no alternate alleles were detected at this position. In the second sample from P007, 614G was detected with a variant allele frequency of 6.1%, but the read depth at this locus (82X) was insufficient to reach our QC standards.

**Table 3.** Frequent non-synonymous variants observed in ≥15 samples (n=47).

| Variant | AF Range | # Patients | # Samples | ORF: Effect |
| --- | --- | --- | --- | --- |
| C17747T | 0.98 - 1 | 11 | 24 | ORF1ab: P5828L; helicase: P504L |
| A17858G | 0.98 - 1 | 11 | 24 | ORF1ab: Y5865C; helicase: Y541C |
| T28144C | 0.02 - 1 | 12 | 23 | ORF8: L84S |
| A23403G | 0.99 - 1 | 9 | 19 | S: D614G |
| C14408T | 0.02 - 1 | 10 | 17 | ORF1ab: P4715L; RdRp: P323L |
| G25563T | 0.03 - 1 | 8 | 15 | ORF3a: Q57H |
| C1059T | 0.95 - 1 | 7 | 15 | ORF1a: T265I; nsp2: T85I |

All variants called had at least 10 reads of support for the alternate allele. For the three variants with large ranges in allele frequency (T28144C, C14408T, G14408T), $\leq 3$ outlier samples with variant AFs below 0.1 were present. When these samples are excluded, minimum AF increases to $\geq 0.99$ .

### Variants exhibiting intra-host evolution are limited in prediction of future global consensus changes and highlight SARS-CoV-2 antibody evasion

We further examined variants that underwent a maximum allele frequency change of ≥20% across timepoints within each patient. The derived alleles for all 25 non-synonymous amino acid changes meeting this criteria in our dataset were also observed among consensus genomes deposited in GISAID [16] by April 2021 (range: 1-2171, mean: 467.8, median: 86.5). Only 8 variants exhibited a maximum allele frequency change of ≥40%. The derived alleles for these variants were present in only 1-1751 GISAID sequences, which at the time of analysis represented a mere 0.0001-0.17% of all sequences deposited in GISAID (Fig 3A). Though the number of consensus sequences with these derived alleles was relatively low, these sequences were diverse with respect to collection date and geographic origin (Fig 3B).

**Fig 3.**
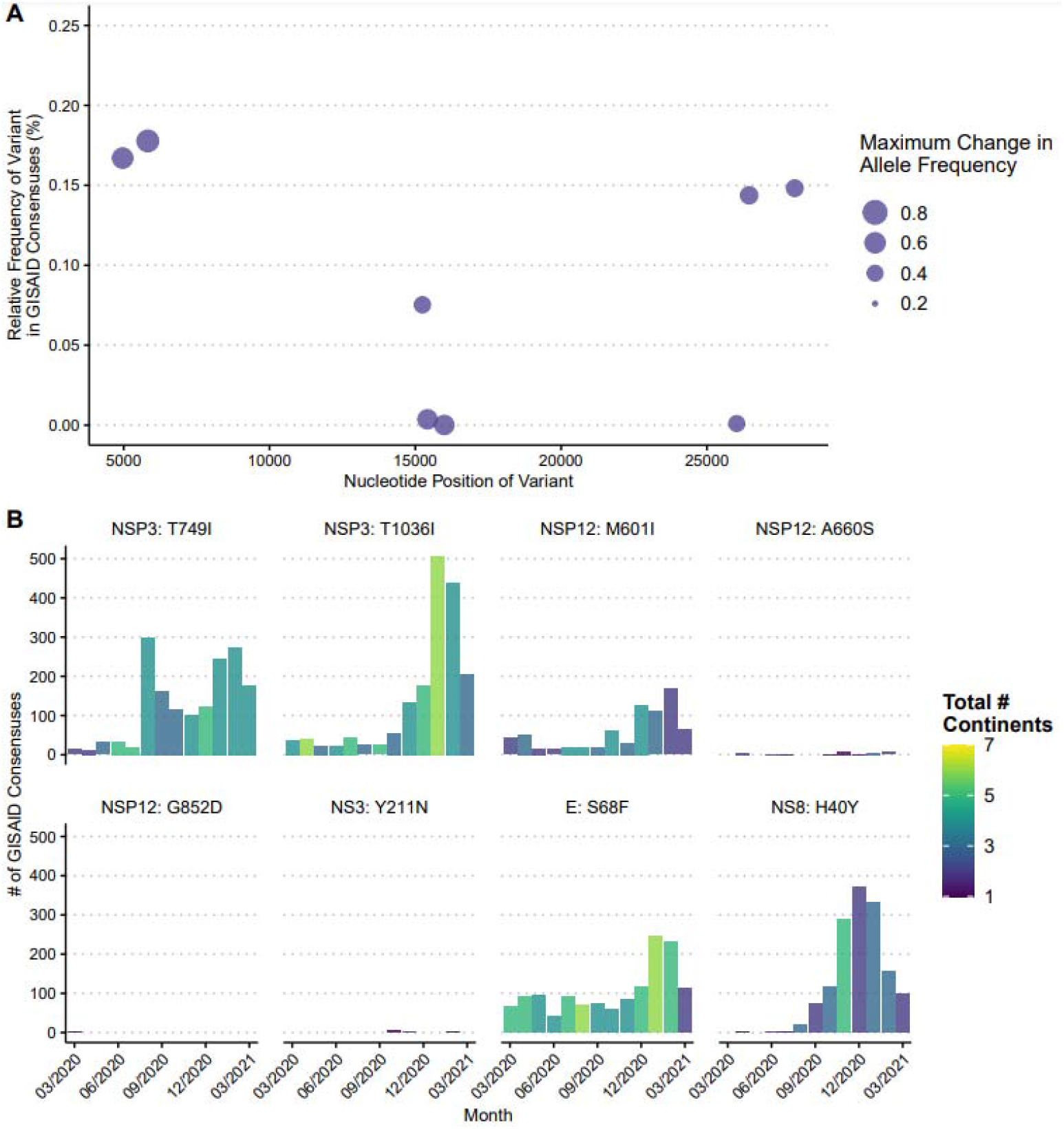
Variants that exhibit ≥40% maximum change in allele frequency in the individuals profiled here in summer 2020 show limited ability to predict future GISAID consensus sequences as of April 2021. A) Relative frequencies of the derived allele found in GISAID consensuses across the genome. Dots represent each unique variant with size indicating the maximum intra-host change in allele frequency found in our study. B) Number of GISAID consensuses with the derived allele for each variant. Height of vertical bars represents the total number of consensuses with the derived allele collected for each month from March 2020 to March 2021 and bar color represents the number of continents of origin for these consensuses.

Three variants with a ≥20% within-host change in allele frequency localized to the spike glycoprotein (Fig 4A). None of these had an allele frequency change of ≥50%. In patient P016, a S:143-145 6-nucleotide deletion was observed at an allele frequency of 20.7% in patient sample 2, but was not observed in samples collected 3 days prior and 3 days later. This patient was immunocompetent and not receiving any COVID-19 treatment. Interestingly, numerous other deletions arose at low frequencies in this patient, with the largest number present at day 0 (Fig 4B). The most prevalent deletion variant in the day 0 sample (collected 16 days after initial symptom onset) was S:Δ141-144 at 1.87% allele frequency. This deletion was the second most common variant in the day 3 sample at 1.2% allele frequency, but was not observed at all in the day 6 sample. Deletions in this region, including S:Δ141-144, have previously been observed in chronically infected immunocompromised patients, and some are associated with escape from NTD-specific neutralizing antibodies or polyclonal sera [7–10,33].

**Fig 4.**
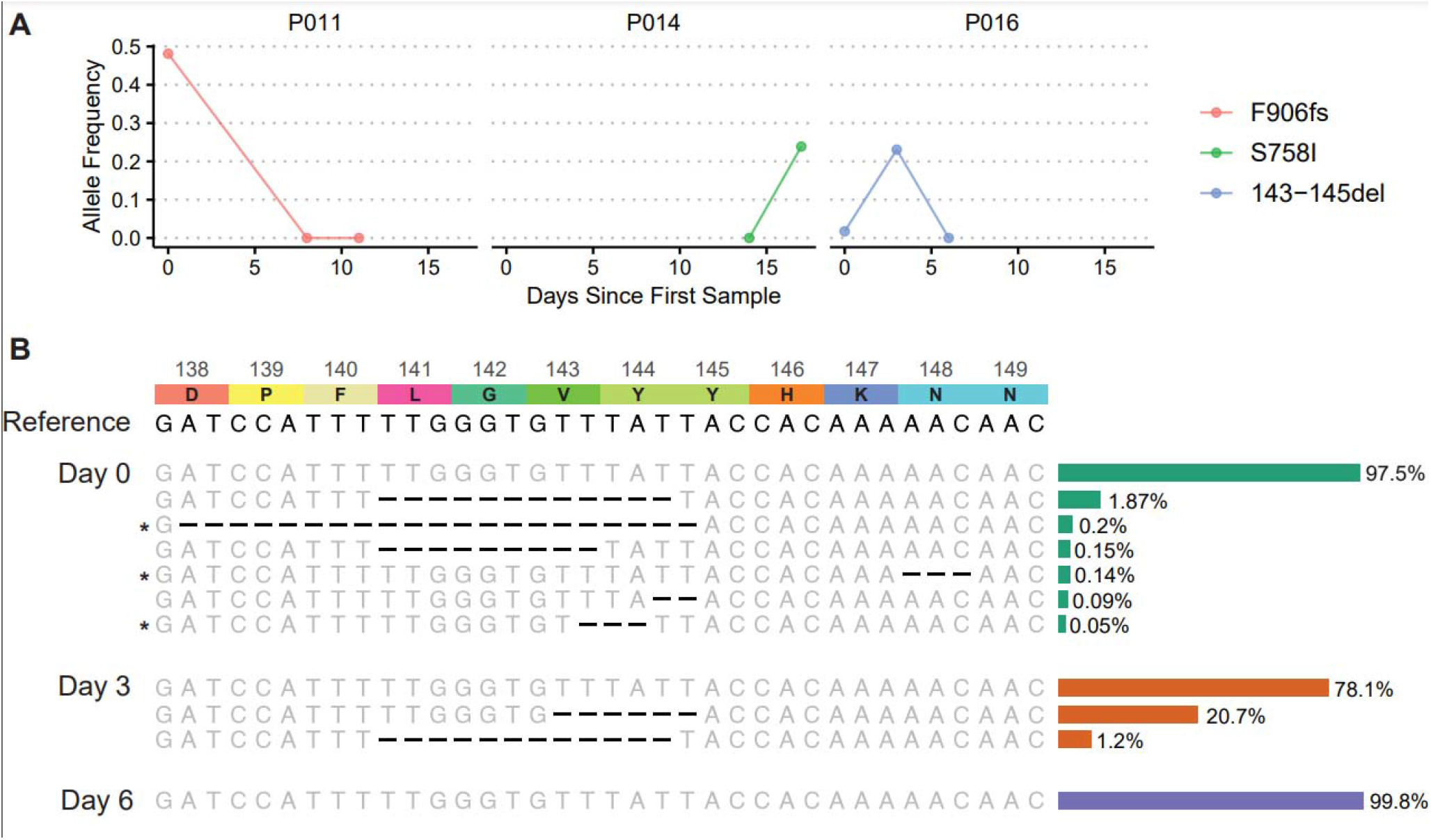
Variants that exhibit intra-host evolution in the spike protein across all patients. A) All non-synonymous variants located in the spike protein with a ≥20% change in allele frequency among timepoints for any patient. B) Enumeration of deletions that arose between residues 138-149 of the spike protein in P016 reveals a rapidly changing complement of low frequency alleles present over a 6-day period. The reference nucleotide sequence (NC_045512) is located at the top of the sequence alignment. Above the reference is the corresponding amino acid sequence with associated residue numbers. Alleles that match the reference are in gray, and deletions are shown in black. Sequences marked with an asterisk (*) may not have deletions depicted in the correct location due to uncertainty of alignment. To the right of the sequence alignment is a bar graph showing the square root of the relative frequency of each variant, for visualization purposes.

### Longitudinal RNAseq analysis illustrates loss of ciliated epithelium during infection

For samples that were sequenced metagenomically, we pseudo-aligned reads to the human transcriptome to perform differential expression analysis comparing initial (t = 0) timepoints to later timepoints. Samples with more than 900,000 pseudo-aligned reads (n = 7 initial, 3 later timepoints) were included in the analysis to determine variation in host gene expression over time. We observed a dramatic downregulation of several cytoskeletal genes, particularly dynein heavy chain (*DNAH 2, 3, 5, 6, 7, 9, 10, 11, 12*), as well as *WDR*s, *MAP1A*, and others (Figure 4A, Supplementary Table 3). Gene Ontology analysis (Figure 4B) confirmed that downregulated genes are involved in biological processes associated with microtubule-based motility. This is consistent with the death of ciliated epithelial cells, which are enriched for transcripts encoding microtubule transport machinery [34], following SARS-CoV-2 infection. We observed upregulation of some actin cytoskeleton-related transcripts like VAV1, VASP, and RhoF [35]. We also observed downregulation of several interferon-stimulated genes, but these did not reach statistical significance in this small sample set.

### Metagenomic analysis shows high levels of clinically relevant bacteria in three samples

We used a previously described metagenomic pipeline (CLOMP [23]) to perform taxonomic assignment of preprocessed reads. We excluded samples that had fewer than 10,000 reads after trimming and any samples that underwent enrichment via probe-capture or amplicon-sequencing for SARS-CoV-2. A total of 24 samples from 11 patients were included in further analysis (Figure 5A, Supplementary Figure 3). No viruses aside from SARS-related coronaviruses met the required cutoffs to be classified as co-infections (see Methods). Multiple samples had detectable numbers of bacterial reads, in particular *Staphylococcus aureus* and *Moraxella catarrhalis*. Samples from two patients (P004, P005) had high detectable levels (>100 RPM) of both bacteria at one or more time point(s) (Figure 5B). In patient P006, who had paired nasopharyngeal (NP) and oropharyngeal (OP) swabs collected at the same time point, we detected *Capnocytophaga gingivalis, Capnocytophaga leadbetteri*, and *Streptococcus parasanguinis* in the OP swab at 21,960, 9,475, and 4,476 RPM, respectively. All three species of bacteria commonly colonize the oropharynx. In contrast, in the NP swab, the predominant species of bacteria was a common skin colonizer, *Cutibacterium acnes*, at 824 RPM. In P010, we found a large number of reads corresponding to *Corynebacterium* spp. at one time point. Upon review of medical records, we found no mention of bacterial co-infections or of positive bacterial cultures in the charts for P004 or P006. P005 had a nares culture that grew methicillin-resistant *Staphylococcus aureus* three months prior to SARS-CoV-2 infection.

**Fig 5.**
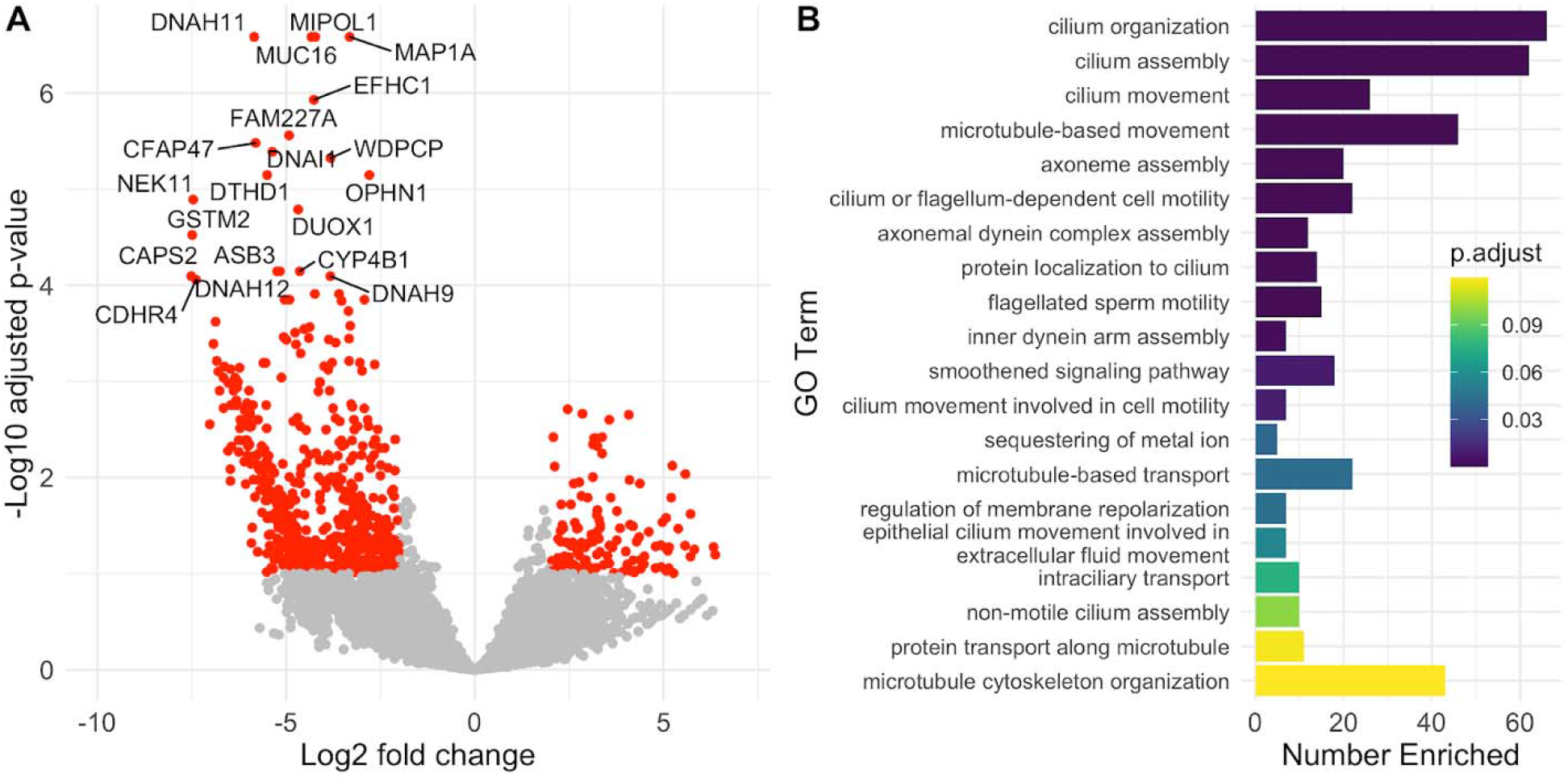
Differentially expressed genes during SARS-CoV-2 infection. A) Twenty differentially expressed genes with lowest adjusted p-value. Fold changes are of later samples relative to initial samples. Genes highlighted in red have a log2 fold change >2 and an adjusted p-value <0.1. B) Gene Ontology analysis reveals that differentially expressed genes are significantly enriched in biological processes related to microtubule-based motility. The twenty biological processes with the lowest adjusted p-values are shown. The length of the horizontal bars corresponds to the number of DE genes in each GO category (“Number Enriched). Bar color corresponds to the adjusted p-value for enrichment of DE genes in each pathway.

**Fig 6.**
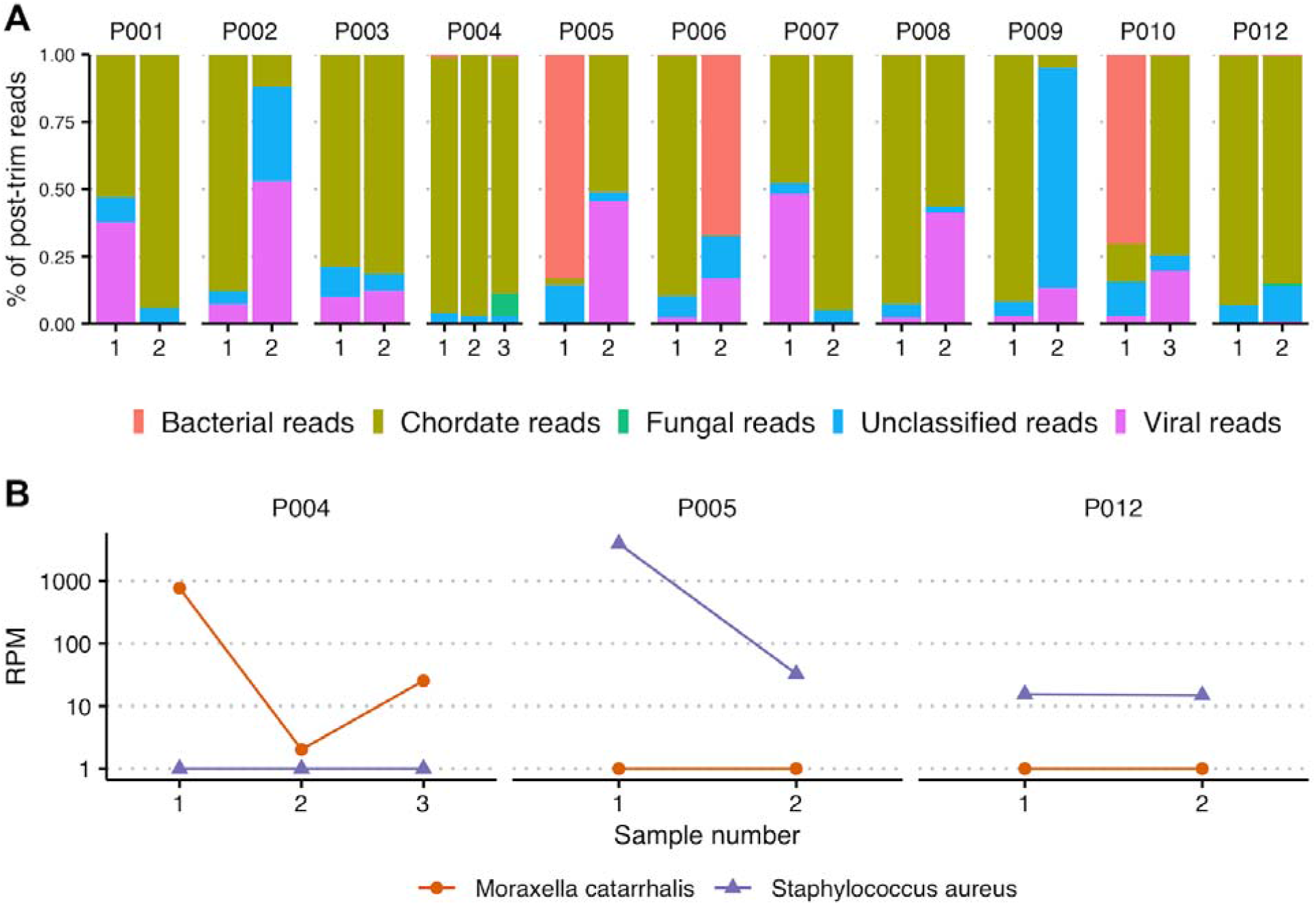
Dynamics of metagenomically classified pathogens. A) Summary of read classifications from CLOMP. Individual bars represent unique samples collected from each patient. Colors correspond to different taxonomic classifications. Reads mapping to SARS-CoV-2 are included in the “Viral reads.” B) Longitudinal RPM values for *Moraxella catarrhalis* and *Staphylococcus aureus* detected by metagenomic analysis.

## Discussion

In this study, we performed high-throughput sequencing of longitudinal clinical specimens that were positive for SARS-CoV-2 by RT-PCR. Most samples were sequenced using a metagenomic approach, which enabled us to simultaneously derive information about viral evolution, host transcription, and the presence of other organisms within patient samples.

We showed that although the viral consensus sequence remains largely unchanged over the course of infection, there is a relative abundance of genome-wide low-frequency variants. Similar to other studies, we saw a wide range in the number of variants detected across samples [36] and distribution of variants across the genome, though some positions appeared to be more prone to variation [2,36]. Studies of SARS-CoV-2 and other respiratory viruses [1,6,7,37] have demonstrated the transmission of minor variants and the role of these population bottlenecks on viral evolution, underscoring the importance of studying within-host viral variation. All variants demonstrating significant longitudinal evolution in our sample set collected March-September 2020 have been observed in consensus sequences from around the globe [16], albeit at relatively low prevalence.

In one patient, we observed rapid turnover of multiple deletion variants in the N-terminal domain of the spike glycoprotein, which has previously been seen in persistent infection in immunocompromised individuals and has been associated with viral escape of neutralizing antibodies [7,9,33,38]. Deletions in the NTD are of particular significance due to their presence in currently circulating lineages of concern. Here we show the emergence of a deletion in this genomic region in an immunocompetent background, suggesting that SARS-CoV-2 can rapidly evolve to escape neutralization within a time span of 1-2 days. It is unclear if the absence of this mutation at day 6 is due to successful clearance of the NTD variant or lack of detection of the minor allele associated with lower copy numbers. In addition, while some of these low frequency deletion alleles have been previously shown to arise independently in different patients in response to similar selection pressures [7], the presence of multiple low frequency deletion alleles within the same patient may be the product of parallel within-host microevolutionary processes. Notably, we did not find any evolving variants selected for in the RBD, the main target for neutralizing activity of human plasma [39,40].

Individual host factors, such as the immune response and respiratory tract microbiome, may play an important role in viral persistence. In particular, because SARS-CoV-2 infection is slow to resolve, the adaptive immune response could drive within-host viral evolution as variants that can escape T-cell and antibody responses develop. Although we were underpowered to see specific evidence of an adaptive immune response being mounted against SARS-CoV-2 in the nasopharynx, detailed studies evaluating antibody and T-cell receptor repertoire changes throughout the course of infection could shed light on the role of immune pressure in the development of minor variants. Similarly, the relationship between bacterial colonization of the nasopharynx and the development or suppression of inflammation in response to SARS-CoV-2 infection remains poorly understood.

As viral load decreases during recovery, it becomes more challenging to recover viral genomes. As a result, one of the limitations of our study is the variability in sequencing depth across samples and the difficulty in ensuring similar sequencing depth for samples from different time points. We used an amplicon sequencing-based approach described previously [27] to obtain near full-length genomes from low viral load samples (up to Ct values of 36). We also used multiple library preparations and performed re-sequencing to ensure the accuracy of variant calls.

Taken together, our results suggest that low frequency genomic variants emerge in immunocompetent individuals, but that these variants are unlikely to reach fixation. Given the emergence of rapidly spreading variants of concern over the past several months, the limited intra-host evolution observed in our dataset highlights the critical impact that a select few individual intra-host evolutionary events may have on the course of the global pandemic and the need for continual genomic surveillance.

## Data Availability

Consensus sequences were deposited to GISAID, and raw reads to SRA under Bioproject PRJNA610428. Accessions are available in Supplementary Table 1.

## Abbreviations

AF: allele frequency
Ct: cycle threshold
NP: nasopharyngeal
NTD: N-terminal domain
OP: oropharyngeal
RBD: receptor-binding domain
RPM: reads per million
RT-PCR: reverse transcription polymerase chain reaction
SD: standard deviation
UW: University of Washington

## Supporting Information

**S1 Fig.**
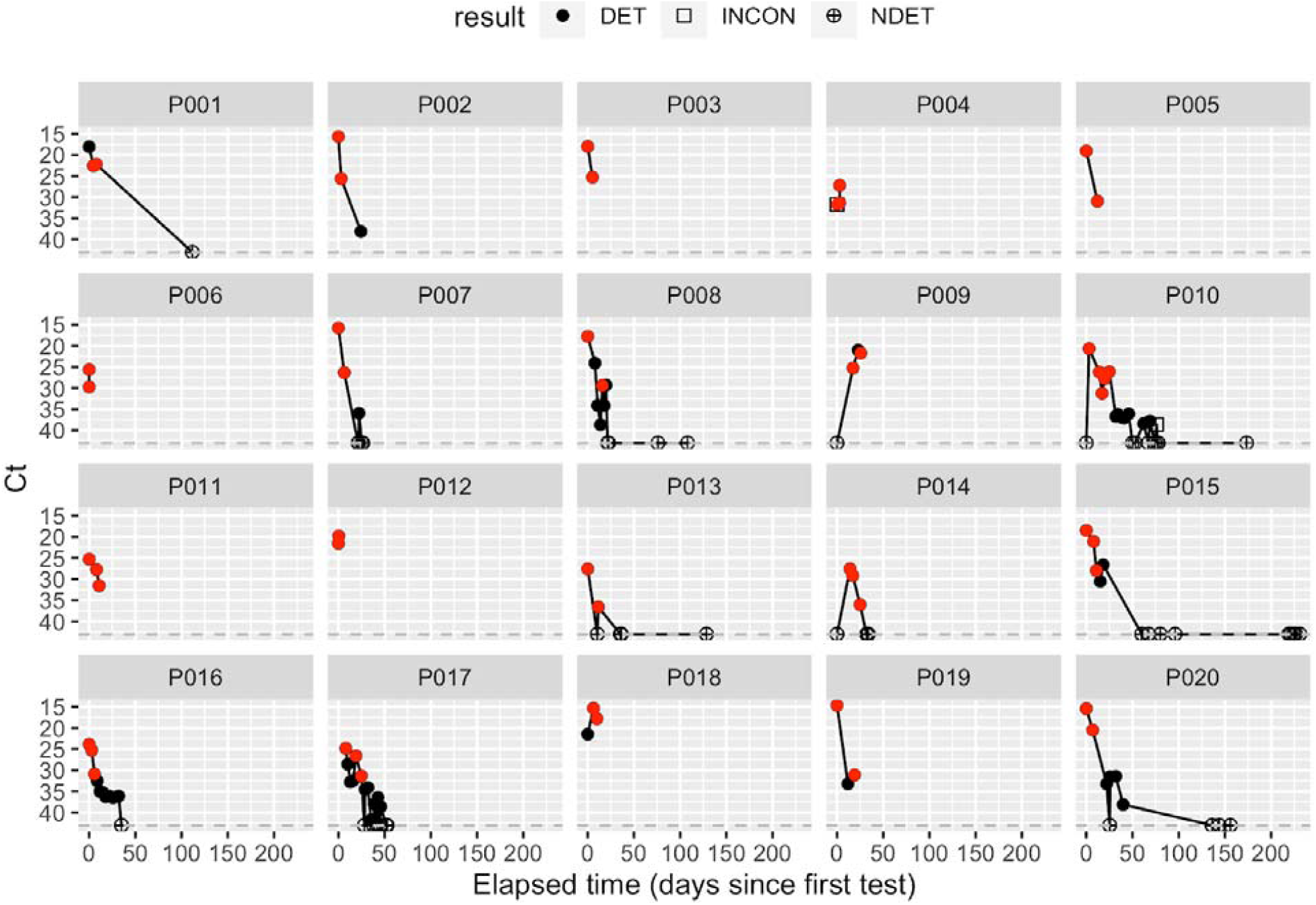
Viral loads for all patients included in the study with samples that returned positive (DET), negative (NDET, Ct >42), and inconclusive (INCON) result by RT-PCR. Red dots represent samples for which we attempted sequencing for this study. Legend above the plot matches shapes of dots to respective RT-PCR results.

**S2 Fig.**
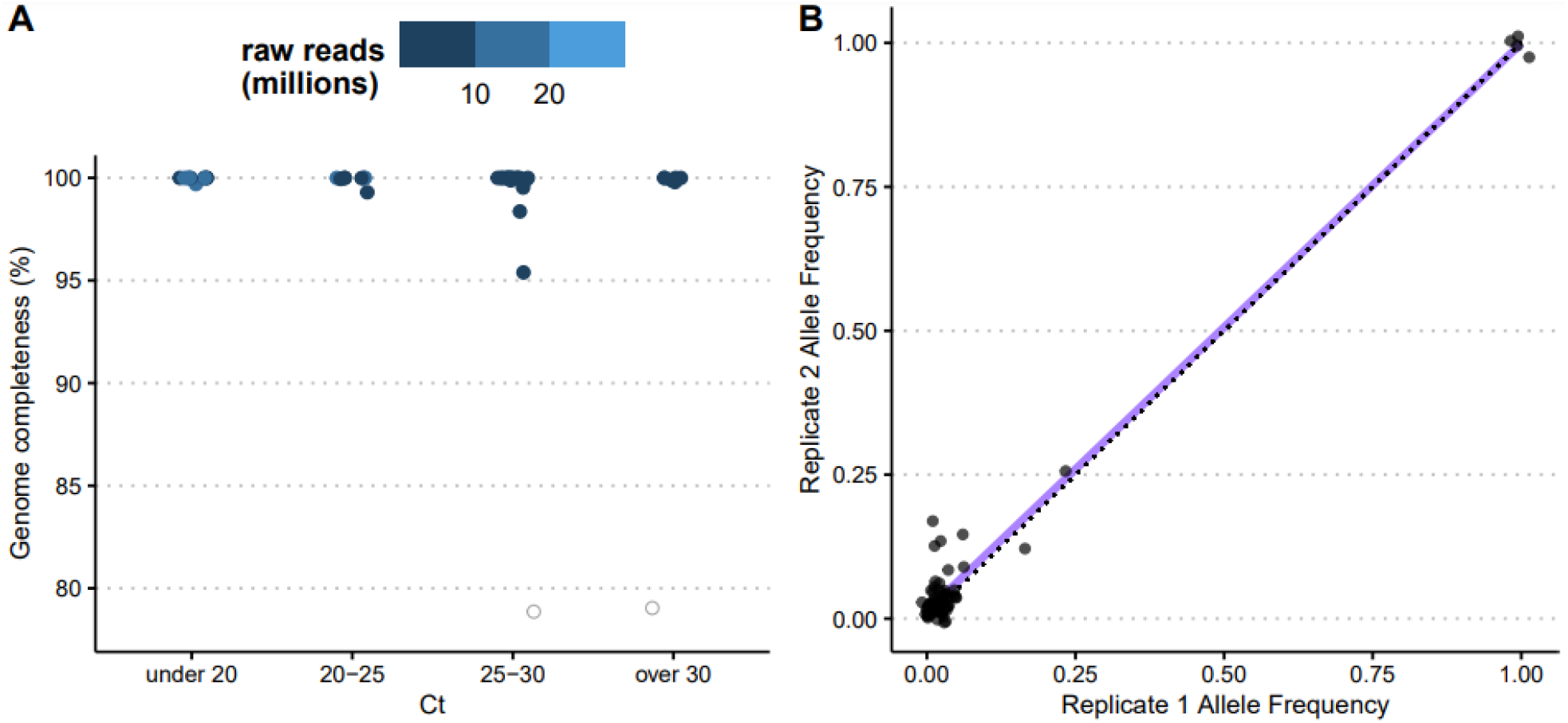
Sequencing quality and reproducibility. (A) Samples were sequenced by metagenomic, probe-capture or amplicon-based approaches. Hollow circles show samples for which a consensus genome could not be recovered. Genome completeness indicates the percentage of non-N nucleotides in consensus sequence (an N was called when there was less than 5X coverage at a site). (B) Comparison of allele frequencies of variants across replicates of the same sample, collapsed across all samples. Each dot represents a variant with ≥50 total depth and ≥10 allelic depth in each replicate. Line of best fit is shown in purple, and x = y dotted line is shown in black.

**S3 Fig.**
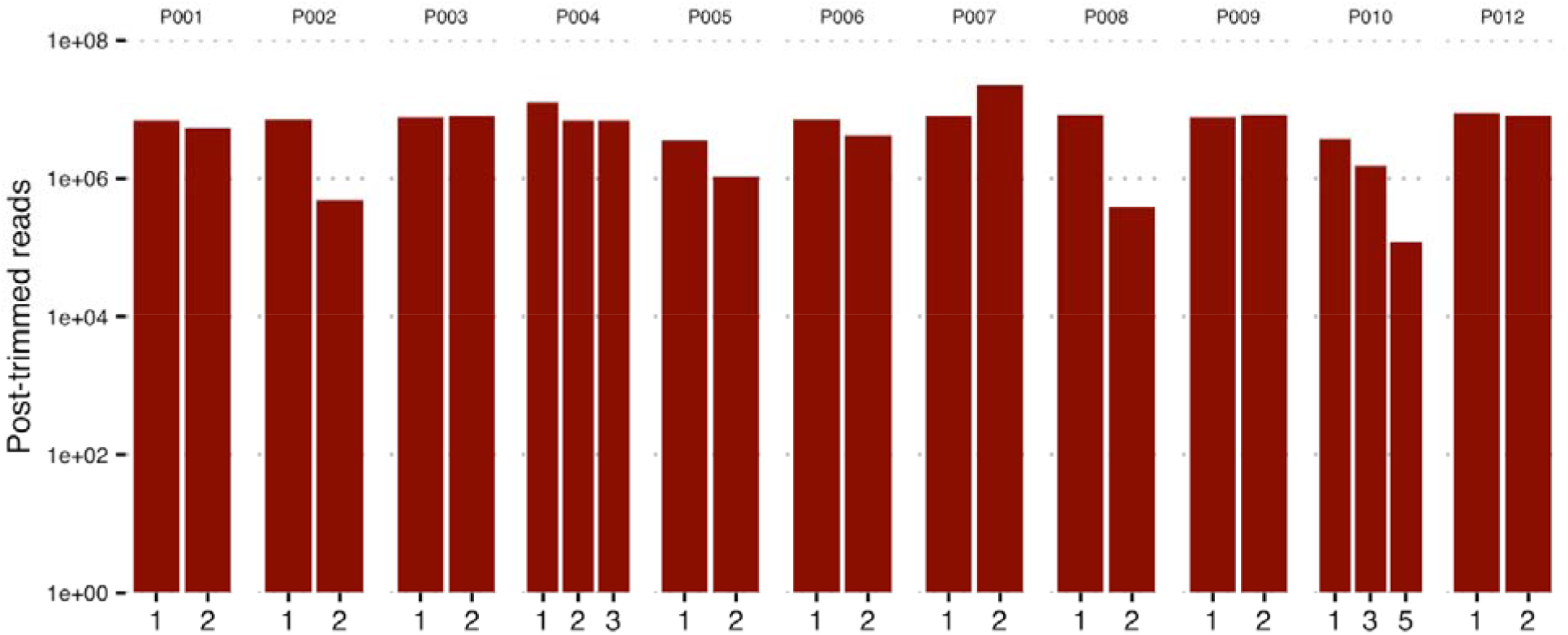
Post-trimmed reads used as input to metagenomic analysis. Samples with under 10,000 reads after trimming, and samples that underwent targeted sequencing (non-metagenomic) were excluded from downstream analysis.

**S1 Table:**
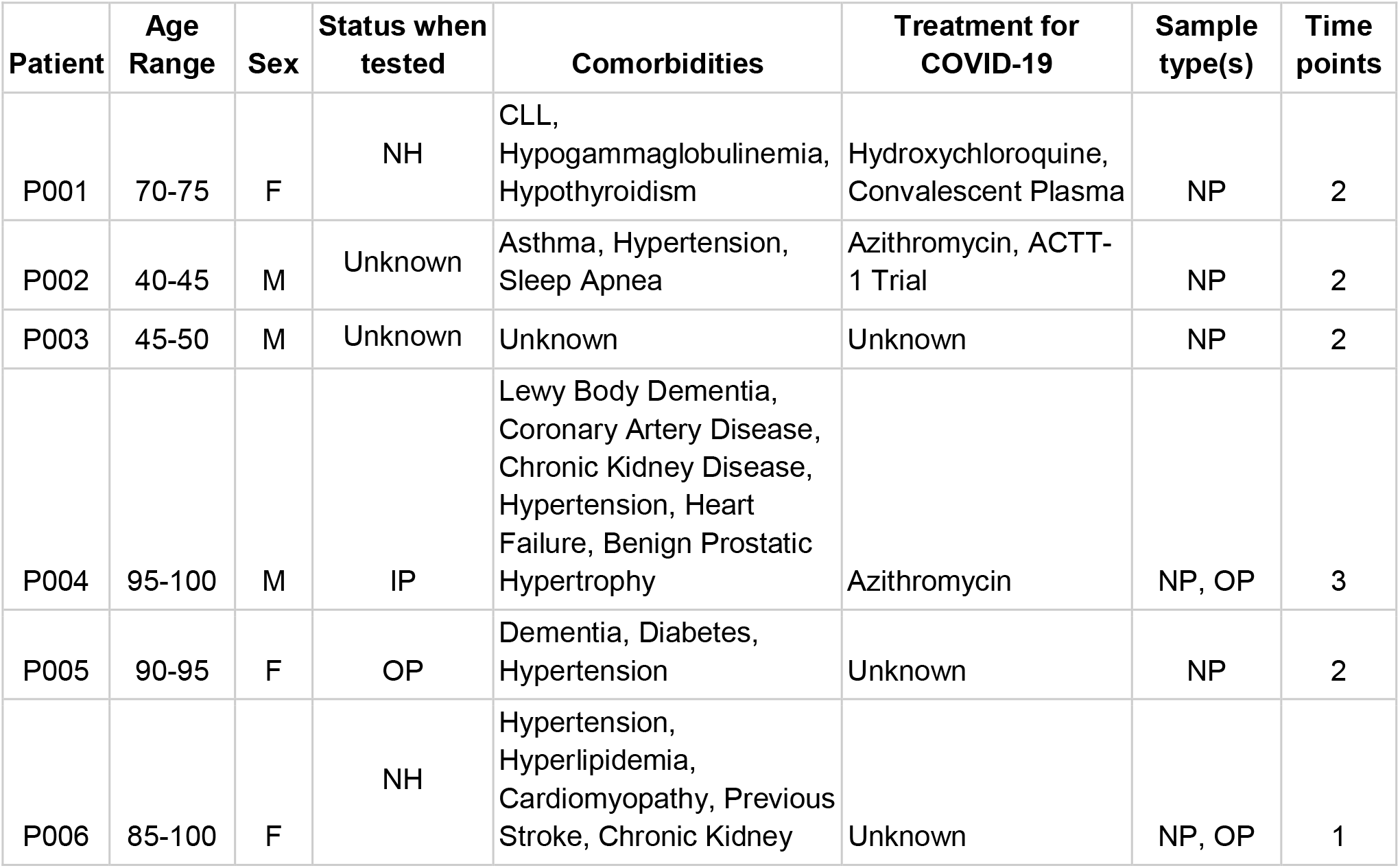
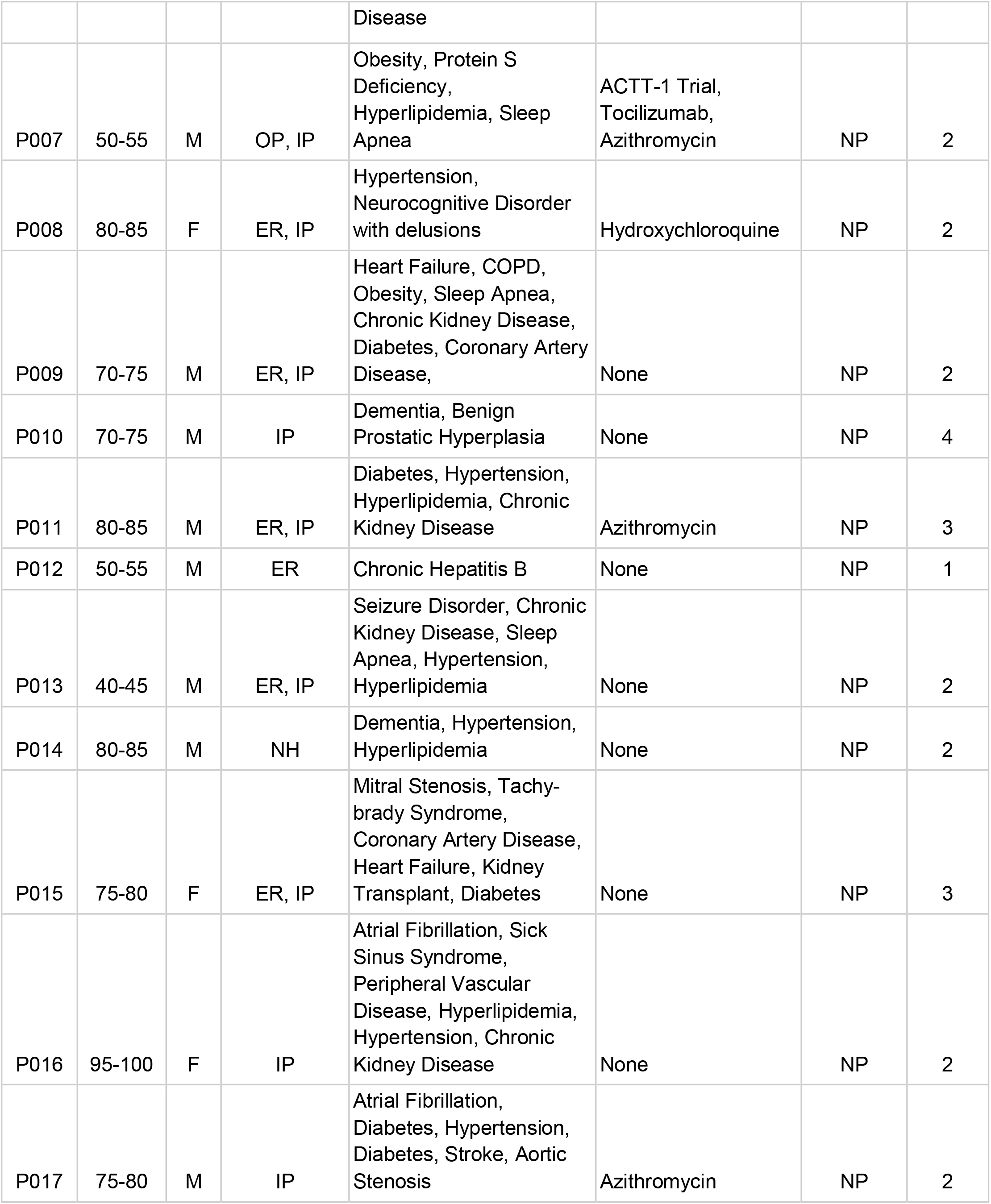
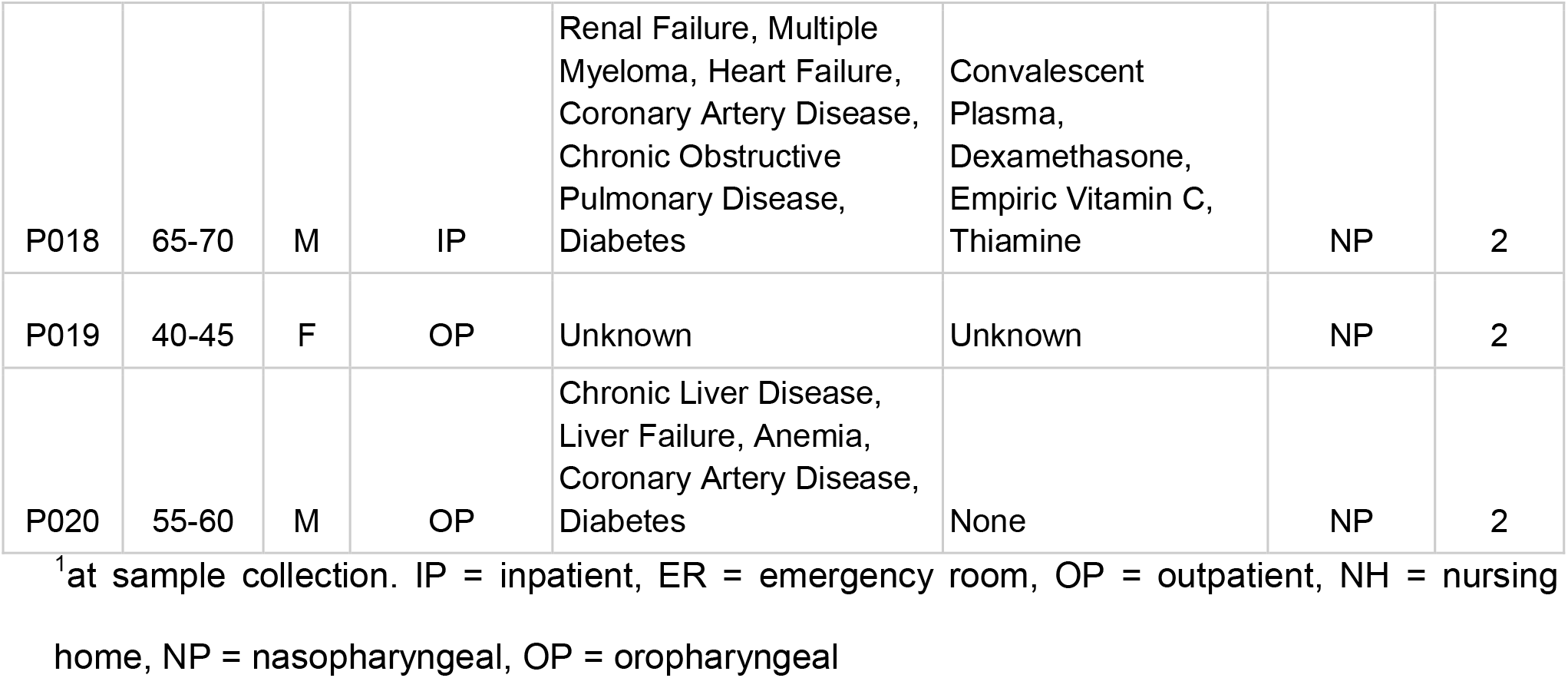
Description of patients included in this study.

**S2 Table.** Data availability (NCBI Bioproject PRJNA610428).

| Patient | Sample # | GISAID accession |
| --- | --- | --- |
| P001 | 1 | EPI_ISL_792092 |
|  | 2 | EPI_ISL_570115 |
| P003 | 1 | EPI_ISL_416643 |
|  | 2 | EPI_ISL_418914 |
| P005 | 1 | EPI_ISL_427186 |
|  | 2 | NA |
| P006 | 1 | EPI_ISL_416454 |
|  | 1 | EPI_ISL_792093 |
| P007 | 1 | EPI_ISL_416448 |
|  | 2 | EPI_ISL_418942 |
| P008 | 1 | EPI_ISL_424228 |
|  | 2 | EPI_ISL_461429 |
| P009 | 1 | EPI_ISL_570975 |
|  | 2 | EPI_ISL_792094 |
| P010 | 1 | EPI_ISL_416717 |
|  | 2 | EPI_ISL_515280 |
|  | 3 | EPI_ISL_792095 |
|  | 4 | EPI_ISL_486097 |
| P011 | 1 | EPI_ISL_570059 |
|  | 2 | EPI_ISL_486102 |
|  | 3 | EPI_ISL_461405 |
| P012 | 1 | EPI_ISL_424190 |
|  | 1 | EPI_ISL_424205 |
| P013 | 1 | EPI_ISL_792096 |
|  | 2 | EPI_ISL_460634 |
| P014 | 1 | EPI_ISL_486103 |
|  | 2 | EPI_ISL_792097 |
| P015 | 1 | EPI_ISL_424262 |
|  | 2 | EPI_ISL_485994 |
|  | 3 | EPI_ISL_792098 |
| P016 | 1 | EPI_ISL_570071 |
|  | 2 | EPI_ISL_825015 |
|  | 3 | EPI_ISL_486110 |
| P017 | 1 | EPI_ISL_792099 |
|  | 2 | EPI_ISL_570078 |
| P018 | 1 | NA |
|  | 2 | NA |
| P019 | 1 | EPI_ISL_570762 |
|  | 2 | EPI_ISL_570779 |
| P020 | 1 | NA |
|  | 2 | EPI_ISL_570503 |

**S3 Table.** Samples used for host gene expression analysis using RNAseq.

| Patient | Sample # | COVID Positivity | Reads Pseudo-aligned to Human Transcriptome |
| --- | --- | --- | --- |
| P001 | 1 | repeat | 914378 |
| P002 | 1 | initial | 1458465 |
| P003 | 1 | initial | 920298 |
| P003 | 2 | repeat | 2115575 |
| P004 | 1 | initial | 11890988 |
| P007 | 2 | repeat | 2450327 |
| P008 | 1 | initial | 1113231 |
| P009 | 1 | initial | 5543628 |
| P015 | 1 | initial | 1544930 |
| P016 | 1 | initial | 1179241 |

**S4 Table.** Clinically relevant taxa.

| Name | Rank | taxID |
| --- | --- | --- |
| Adenoviridae | F | 10508 |
| Human coronavirus HKU1 | S | 290028 |
| Rousettus bat coronavirus HKU9 | S | 694006 |
| Human coronavirus NL63 | S | 277944 |
| Human coronavirus 229E | S | 11137 |
| Severe acute respiratory syndrome-related coronavirus | S | 694009 |
| Enterovirus | G | 12059 |
| Rhinovirus C | S | 463676 |
| Propionibacterium phage PacnesP2 | S | 1983621 |
| Propionibacterium phage pa33 | S | 2079406 |
| Propionibacterium phage Aquarius | S | 2041558 |
| Propionibacterium phage Moyashi | S | 1654781 |
| Propionibacterium phage PHL055N00 | S | 1500802 |
| Propionibacterium virus Pacnes201215 | S | 1982270 |
| Propionibacterium phage Pacnes 2012-15 | - | 1498188 |
| Streptococcus virus MS1 | S | 1962672 |
| environmental samples | - | 2100420 |
| uncultured Caudovirales phage | S | 2100421 |
| unclassified Papillomaviridae | - | 333774 |
| Human papillomavirus types | - | 173087 |
| Human papillomavirus | S | 10566 |
| Betacoronavirus BtCoV/Rhi_hip/R8-09/SPA/2010 | S | 1346312 |
| Bat Hp-betacoronavirus Zhejiang2013 | S | 2501961 |
| Bat Hp-betacoronavirus/Zhejiang2013 | - | 1541205 |
| Colobus guereza | S | 33548 |
| Influenza A virus | S | 11320 |
| Bordetella pertussis | S | 520 |
| Staphylococcus aureus | S | 1280 |
| Staphylococcus aureus | S | 1280 |
| Haemophilus influenzae | S | 727 |
| Pseudomonas aeruginosa | S | 287 |
| Pseudomonas oleovorans/pseudoalcaligenes group | - | 1232139 |
| Streptococcus pneumoniae | S | 1313 |
| Streptococcus sp. A12 | S | 1759399 |
| Streptococcus pseudopneumoniae | S | 257758 |
| Streptococcus pseudopneumoniae IS7493 | - | 1054460 |
| Streptococcus sp. I-G2 | S | 1156431 |
| Moraxella catarrhalis | S | 480 |

## References

1. Xue KS, Moncla LH, Bedford T, Bloom JD. Within-Host Evolution of Human Influenza Virus. Trends Microbiol. 2018 Sep;26(9):781–93.

2. van Dorp L, Acman M, Richard D, Shaw LP, Ford CE, Ormond L, et al. Emergence of genomic diversity and recurrent mutations in SARS-CoV-2. Infect Genet Evol. 2020 Sep;83:104351.

3. Sashittal P, Luo Y, Peng J, El-Kebir M. Characterization of SARS-CoV-2 viral diversity within and across hosts. BioRxiv [Preprint]. 2020 May [cited 2021 Mar 24]. Available from: http://biorxiv.org/lookup/doi/10.1101/2020.05.07.083410

4. Ramazzotti D, Angaroni F, Maspero D, Gambacorti-Passerini C, Antoniotti M, Graudenzi A, et al. VERSO: A comprehensive framework for the inference of robust phylogenies and the quantification of intra-host genomic diversity of viral samples. Patterns. 2021 Mar;2(3):100212.

5. Rose R, Nolan DJ, Moot S, Feehan A, Cross S, Garcia-Diaz J, et al. Intra-host site-specific polymorphisms of SARS-CoV-2 is consistent across multiple samples and methodologies. MedRxiv [Preprint]. 2020 Apr [cited 2021 Mar 24]. Available from: http://medrxiv.org/lookup/doi/10.1101/2020.04.24.20078691

6. Lythgoe KA, Hall M, Ferretti L, de Cesare M, MacIntyre-Cockett G, Trebes A, et al. SARS-CoV-2 within-host diversity and transmission. Science. 2021 Mar 9;eabg0821.

7. McCarthy KR, Rennick LJ, Nambulli S, Robinson-McCarthy LR, Bain WG, Haidar G, et al. Recurrent deletions in the SARS-CoV-2 spike glycoprotein drive antibody escape. Science. 2021 Mar 12;371(6534):1139–42.

8. Chen L, Zody MC, Mediavilla JR, Cunningham MH, Composto K, Chow KF, et al. Emergence of multiple SARS-CoV-2 antibody escape variants in an immunocompromised host undergoing convalescent plasma treatment [Internet]. Infectious Diseases (except HIV/AIDS); 2021 Apr [cited 2021 Apr 14]. Available from: http://medrxiv.org/lookup/doi/10.1101/2021.04.08.21254791

9. McCallum M, De Marco A, Lempp FA, Tortorici MA, Pinto D, Walls AC, et al. N-terminal domain antigenic mapping reveals a site of vulnerability for SARS-CoV-2. Cell. 2021 Mar;S0092867421003561.

10. Andreano E, Piccini G, Licastro D, Casalino L, Johnson NV, Paciello I, et al. SARS-CoV-2 escape in vitro from a highly neutralizing COVID-19 convalescent plasma. BioRxiv [Preprint]. 2020 Dec [cited 2021 Apr 17]. Available from: http://biorxiv.org/lookup/doi/10.1101/2020.12.28.424451

11. Perchetti GA, Nalla AK, Huang M-L, Zhu H, Wei Y, Stensland L, et al. Validation of SARS-CoV-2 detection across multiple specimen types. J Clin Virol. 2020 Jul;128:104438.

12. Perchetti GA, Sullivan K-W, Pepper G, Huang M-L, Breit N, Mathias P, et al. Pooling of SARS-CoV-2 samples to increase molecular testing throughput. J Clin Virol. 2020 Oct;131:104570.

13. Greninger AL, Zerr DM, Qin X, Adler AL, Sampoleo R, Kuypers JM, et al. Rapid Metagenomic Next-Generation Sequencing during an Investigation of Hospital-Acquired Human Parainfluenza Virus 3 Infections. J Clin Microbiol. 2017 Jan;55(1):177–82.

14. Greninger AL, Roychoudhury P, Xie H, Casto A, Cent A, Pepper G, et al. Ultrasensitive Capture of Human Herpes Simplex Virus Genomes Directly from Clinical Samples Reveals Extraordinarily Limited Evolution in Cell Culture. Fernandez-Sesma a, editor. mSphere. 2018 Jun 13;3(3):e00283–18, /msphere/3/3/mSphere283-18.atom.

15. Addetia A, Lin MJ, Peddu V, Roychoudhury P, Jerome KR, Greninger AL. Sensitive Recovery of Complete SARS-CoV-2 Genomes from Clinical Samples by Use of Swift Biosciences’ SARS-CoV-2 Multiplex Amplicon Sequencing Panel. Dekker JP, editor. J Clin Microbiol. 2020 Dec 17;59(1):JCM.02226-20, e02226–20.

16. Shu Y, McCauley J. GISAID: Global initiative on sharing all influenza data – from vision to reality. Eurosurveillance [Internet]. 2017 Mar 30 [cited 2021 Mar 24];22(13). Available from: https://www.eurosurveillance.org/content/10.2807/1560-7917.ES.2017.22.13.30494

17. Katoh K, Standley DM. MAFFT Multiple Sequence Alignment Software Version 7: Improvements in Performance and Usability. Mol Biol Evol. 2013 Apr 1;30(4):772–80.

18. Hadfield J, Megill C, Bell SM, Huddleston J, Potter B, Callender C, et al. Nextstrain: real-time tracking of pathogen evolution. Kelso J, editor. Bioinformatics. 2018 Dec 1;34(23):4121–3.

19. Bolger AM, Lohse M, Usadel B. Trimmomatic: a flexible trimmer for Illumina sequence data. Bioinformatics. 2014 Aug 1;30(15):2114–20.

20. Bray NL, Pimentel H, Melsted P, Pachter L. Near-optimal probabilistic RNA-seq quantification. Nat Biotechnol. 2016 May;34(5):525–7.

21. Love MI, Huber W, Anders S. Moderated estimation of fold change and dispersion for RNA-seq data with DESeq2. Genome Biol. 2014 Dec;15(12):550.

22. Yu G, Wang L-G, Han Y, He Q-Y. clusterProfiler: an R package for comparing biological themes among gene clusters. Omics J Integr Biol. 2012 May;16(5):284–7.

23. Peddu V, Shean RC, Xie H, Shrestha L, Perchetti GA, Minot SS, et al. Metagenomic Analysis Reveals Clinical SARS-CoV-2 Infection and Bacterial or Viral Superinfection and Colonization. Clin Chem. 2020 Jul 1;66(7):966–72.

24. Breitwieser FP, Salzberg SL. Pavian: interactive analysis of metagenomics data for microbiome studies and pathogen identification. Schwartz R, editor. Bioinformatics. 2020 Feb 15;36(4):1303–4.

25. Nalla AK, Casto AM, Huang M-LW, Perchetti GA, Sampoleo R, Shrestha L, et al. Comparative Performance of SARS-CoV-2 Detection Assays Using Seven Different Primer-Probe Sets and One Assay Kit. McAdam AJ, editor. J Clin Microbiol. 2020 Apr 8;58(6):e00557–20, /jcm/58/6/JCM.00557-20.atom.

26. He X, Lau EHY, Wu P, Deng X, Wang J, Hao X, et al. Temporal dynamics in viral shedding and transmissibility of COVID-19. Nat Med. 2020 May;26(5):672–5.

27. Addetia A, Xie H, Roychoudhury P, Shrestha L, Loprieno M, Huang M-L, et al. Identification of multiple large deletions in ORF7a resulting in in-frame gene fusions in clinical SARS-CoV-2 isolates. J Clin Virol. 2020 Aug;129:104523.

28. Bedford T, Greninger AL, Roychoudhury P, Starita LM, Famulare M, Huang M-L, et al. Cryptic transmission of SARS-CoV-2 in Washington state. Science. 2020 Oct 30;370(6516):571–5.

29. Worobey M, Pekar J, Larsen BB, Nelson MI, Hill V, Joy JB, et al. The emergence of SARS-CoV-2 in Europe and North America. Science. 2020 Oct 30;370(6516):564–70.

30. Korber B, Fischer WM, Gnanakaran S, Yoon H, Theiler J, Abfalterer W, et al. Tracking Changes in SARS-CoV-2 Spike: Evidence that D614G Increases Infectivity of the COVID-19 Virus. Cell. 2020 Aug;182(4):812-827.e19.

31. Müller NF, Wagner C, Frazar CD, Roychoudhury P, Lee J, Moncla LH, et al. Viral genomes reveal patterns of the SARS-CoV-2 outbreak in Washington State. MedRxiv [Preprint]. 2020 Sep [cited 2021 Mar 24]. Available from: http://medrxiv.org/lookup/doi/10.1101/2020.09.30.20204230

32. Yurkovetskiy L, Wang X, Pascal KE, Tomkins-Tinch C, Nyalile TP, Wang Y, et al. Structural and Functional Analysis of the D614G SARS-CoV-2 Spike Protein Variant. Cell. 2020 Oct;183(3):739-751.e8.

33. Avanzato VA, Matson MJ, Seifert SN, Pryce R, Williamson BN, Anzick SL, et al. Case Study: Prolonged Infectious SARS-CoV-2 Shedding from an Asymptomatic Immunocompromised Individual with Cancer. Cell. 2020 Dec;183(7):1901-1912.e9.

34. Maiti AK, Mattéi M-G, Jorissen M, Volz A, Zeigler A, Bouvagnet P. Identification, tissue specific expression, and chromosomal localisation of several human dynein heavy chain genes. Eur J Hum Genet. 2000 Dec;8(12):923–32.

35. Zhu N, Wang W, Liu Z, Liang C, Wang W, Ye F, et al. Morphogenesis and cytopathic effect of SARS-CoV-2 infection in human airway epithelial cells. Nat Commun. 2020 Dec;11(1):3910.

36. Shen Z, Xiao Y, Kang L, Ma W, Shi L, Zhang L, et al. Genomic Diversity of Severe Acute Respiratory Syndrome–Coronavirus 2 in Patients With Coronavirus Disease 2019. Clin Infect Dis. 2020 Jul 28;71(15):713–20.

37. Lythgoe KA, Hall M, Ferretti L, de Cesare M, MacIntyre-Cockett G, Trebes A, et al. Within-host genomics of SARS-CoV-2. BioRxiv [Preprint]. 2020 May [cited 2021 Mar 24]. Available from: http://biorxiv.org/lookup/doi/10.1101/2020.05.28.118992

38. Choi B, Choudhary MC, Regan J, Sparks JA, Padera RF, Qiu X, et al. Persistence and Evolution of SARS-CoV-2 in an Immunocompromised Host. N Engl J Med. 2020 Dec 3;383(23):2291–3.

39. Greaney AJ, Loes AN, Crawford KHD, Starr TN, Malone KD, Chu HY, et al. Comprehensive mapping of mutations in the SARS-CoV-2 receptor-binding domain that affect recognition by polyclonal human plasma antibodies. Cell Host Microbe. 2021 Mar;29(3):463-476.e6.

40. Piccoli L, Park Y-J, Tortorici MA, Czudnochowski N, Walls AC, Beltramello M, et al. Mapping Neutralizing and Immunodominant Sites on the SARS-CoV-2 Spike Receptor-Binding Domain by Structure-Guided High-Resolution Serology. Cell. 2020 Nov;183(4):1024-1042.e21.

